# Common pathogenic mechanisms for COVID-19 and differentiated thyroid cancer: a proteomic analysis

**DOI:** 10.1101/2024.09.28.24314528

**Authors:** Hanqing Liu, Jiaxi Wang, Dan Yang, Chuang Chen

## Abstract

**Background:** Previous studies have proved that viral infection might have potential associations with differentiated thyroid cancer. COVID-19 has spread to hundreds of millions of individuals since Dec 2019. This study aimed to investigate the common pathogenic mechanisms of differentiated thyroid cancer and SARS-CoV-2 infection in thyroid tissues.

**Methods:** The proteomic profiles of COVID-19 and differentiated thyroid cancer were downloaded from iProx database and were analyzed for differentially expressed proteins. After the common proteins were identified using the Venn diagram, gene ontology and pathway enrichment analysis were performed. Subsequently, a protein-protein interaction network was constructed and hub genes were identified with eight algorithms. The diagnostic performance of hub genes was tested with the receiver operator characteristics curve. The associations between hub genes and diseases were evaluated with the Comparative Toxicogenomics Database.

**Results:** Forty-nine common differentially expressed proteins were identified. Functional analysis revealed that the metabolism and transport of lipid and cholesterol and coagulation process were the main common biological activities affected by the two diseases. In addition, twelve genes, including AGT, AHSG, APOA2, APOM, C3, GC, ITIH2, KNG1, SERPINA1, SERPINC1, TF and TTR, were identified as the hub genes. These genes could serve as diagnostic markers for COVID-19 and differentiated thyroid cancers.

**Conclusion:** The current study revealed common pathogenic mechanisms of COVID-19 and differentiated thyroid cancer. The concomitant infection of SARS-CoV-2 might exert adverse effects on patients with thyroid cancers.

## 1 Introduction

Thyroid cancer is the most common endocrine malignancy all over the world (1). Its incidence has doubled since the mid-1990s (2, 3). Although thyroid cancer consists of several histopathological types, papillary thyroid cancer (PTC) and follicular thyroid cancer (FTC) account for more than 80% of total cases. PTC and FTC are also categorized as differentiated thyroid cancer (DTC) in contrast with poorly differentiated and anaplastic thyroid cancers. Notably, DTC contributes to the majority of incidence elevation (4). While the incidence of other histopathological subtypes remains almost unaltered, the incidence of DTC increased by 300% since 1975 (5). The elevation may be explained by both increasing exposure to risk factors and the application of ultrasound devices with high resolution.

The risk factors for thyroid cancers are various, including radiation, iodine deficiency, genetic susceptibility, etc. More recently, the association between thyroid cancer and viral infection has been noticed. Epstein-Barr virus is a well-known human tumor virus associated with multiple malignancies, including nasopharyngeal carcinoma, Hodgkin’s lymphoma and Burkitt’s lymphoma. A retrospective clinical study using *in situ* hybridization showed that the detection of Epstein-Barr virus (EBV) was significantly higher in thyroid cancer tissues compared with paired adjacent normal tissues (6). Similar results were verified with serologic assays in another study (7). In addition, the potential link between hepatitis C virus and thyroid cancer has been revealed. A higher prevalence of chronic hepatitis was found in patients with thyroid cancers (8). Similar phenomena were also found in patients infected with hepatitis B virus (9, 10), human erythrovirus B19 (11, 12), cytomegalovirus (13), and human T-cell lymphotropic virus (14). Of interest, these viruses are also related to autoimmune thyroiditis (15). The carcinogenetic mechanism of these viruses, if exists, is most likely to be chronic inflammation. Some virus proteins, such as latent membrane protein released by EBV, can exert effects on cell cycle-related intracellular proteins. Inferably, viral oncogene may also be a potential mechanism in the carcinogenesis of thyroid cancer.

Since the first case was reported in Wuhan, China in Dec 2019, coronavirus disease 2019 (COVID-19) has spread to 228 countries and territories. More than six hundred million cases were confirmed and its mortality is approximately 1% (16). Undoubtedly, the real incidence is much higher than the reported number since nucleic acid tests and antigen assays are not performed in all suspected patients. The pathogen of COVID-19 is severe acute respiratory syndrome coronavirus 2 (SARS-CoV-2). The new coronavirus can invade peripheral tissue cells via the angiotensin-converting enzyme 2 (ACE2) and cause multiple organ dysfunctions, including lung, kidney and heart (17). Direct viral infection, cytokine storm, down-regulation of ACE2 and endothelial cell damage are supposed to be the pathogenic mechanisms (18–20).

Of interest, current evidences suggest that potential linkages may exist between thyroid cancer and COVID-19. A multicenter cohort study in Los Angeles revealed a positive correlation between comorbid conditions and risk of hospitalization from SARS-CoV-2 infection in thyroid cancer patients (21). Another retrospective study revealed a small but significant elevation of DTC after the outbreak of the pandemic, although the results were hard to explain and confounding factors needed to be explored (22). Autoimmune thyroid diseases are known to increase the risk of thyroid cancer. Previous reports have shown that the relapse or new diagnosis of autoimmune thyroid diseases could be triggered within one month after SARS-CoV-2 infection (23). Some researchers have assumed that thyroid inflammation could be induced by COVID-19-related immune response (24). Since the ACE2 expression levels are higher in thyroid tissue in comparison with lung (25), the coronavirus might damage normal thyroid tissues and subsequently induce carcinogenesis. Currently, little is known about the association between COVID-19 and DTC from a molecular aspect.

The aim of this study is to evaluate the potential common molecular mechanisms for DTC and COVID-19 using bioinformatic tools.

## 2 Materials and methods

### 2.1 Data acquisition

Two proteomic expression profiles were selected and downloaded from iProX database (Figure 1) (26). iProX database is a public platform for collecting and sharing proteomics datasets. The two proteomic datasets were both released by Guo’s lab at Westlake University (27, 28). The tissue biopsy samples were processed using the pressure cycling technique protocol to separate small amounts of components. Subsequently, data-independent acquisition mass spectrometry was used to analyze the prepared samples at high throughput. The IPX0002393000 dataset displayed the proteomic profiles of various organs in patients with COVID-19 (Table 1). An amount of 15 thyroid samples from the autopsy of infected patients and 15 from non-infected patients were extracted. The IPX0001444000 dataset included 1161 samples of various thyroid diseases, including differentiated thyroid cancer (DTC), benign thyroid diseases and normal thyroid tissues (NT). Two hundred and twenty-three samples of differentiated thyroid cancer or normal thyroid tissues in the discovery subgroup were collected for subsequent analysis.

**Figure 1.**
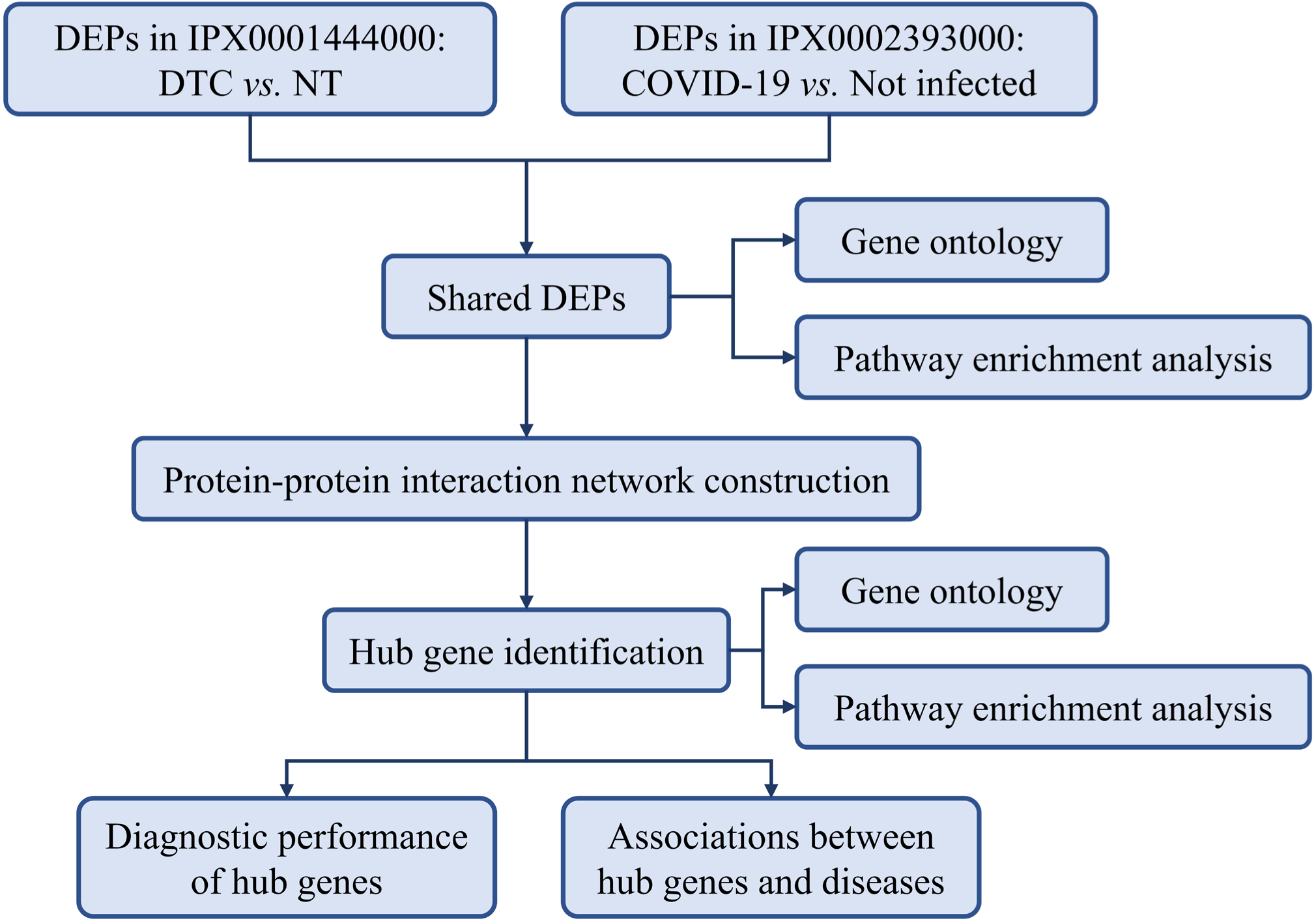
The flow chart of the study. Abbreviations: COVID-19, coronavirus disease 2019; DEP, differentially expressed protein; DTC, differentiated thyroid cancer; NT normal thyroid tissue.

**Table 1.** The summary of the included datasets.

| Series number | Organism | Sample type | Diseases | Release date | Number of samples<br>(used/total) | Groups |
| --- | --- | --- | --- | --- | --- | --- |
| IPX0001444000 | Human | FFPE, FNAB | PTC, FTC, benign thyroid diseases,<br>normal thyroid tissues | Sept 6, 2022 | 223/1161 <sup>a</sup> | FTC/PTC/normal:<br>64/119/40 |
| IPX0002393000 | Human | FFPE | COVID-19, normal thyroid tissues | Dec 11, 2020 | 30/218 | COVID-19/normal: 15/15 |

Note: The two datasets were provided by Westlake laboratory of life sciences and biomedicine, key laboratory of structural biology of Zhejiang province, school of life sciences, Westlake University.
Abbreviations: COVID-19, coronavirus disease 2019; FFPE, formalin-fixed paraffin-embedded tissue; FNAB, fine-needle aspiration biopsy; FTC, follicular thyroid cancer; PTC, papillary thyroid cancer
<sup>a</sup> A total of 1133 patients were involved in the dataset. More than one samples could be collected from different nodules of one patient.

### 2.2 Identification of DEPs

Expression profiles were inputted into R project (v4.2.0). The clinical information was collected from the two datasets for sample classification. Samples of irrelevant diseases were excluded from the expression matrices. Missing data were imputed using the k-nearest neighbors algorithm in *DMwR2* package. Since technical replicates for each sample were set and batch effects were minimized at the batch design level, the robust multi-array normalization was not conducted so as not to eliminate the true difference (Sup figure 1). UniProtKB AC/IDs were converted to general gene symbols via the online tool UniProt (29). When multiple protein IDs corresponded to one gene symbol, the medians were calculated. The Bayesian method in the *limma* R package was used to identify the differentially expressed proteins (DEPs) in two comparisons (COVID-19 *vs.* not infected, and DTC *vs.* NT). *P*-values were adjusted using the Benjamini-Hochberg method. Fold-changes (FCs) were calculated with the false-discovery rate procedure. DTC-DEGs with |log2-FC| >1 and *p* <0.01 were selected. COVID-19-DEGs were filtered according to the criteria |log2-FC| >0.5 and *p* <0.01. The Venn diagram of DEGs was constructed with SangerBox (v3.0) (30) to identify the overlapping DEPs. Heatmaps were constructed to visualize the common DEPs in the two expression profiles.

### 2.3 Gene ontology and pathway enrichment analysis

Gene ontology (GO) annotation and pathway analysis were performed with three online tools, namely, DAVID (v2022q3), AmiGO (v2.5.17) and Reactome (v83) (31–33). The GO enrichment analysis included three categories: biological process (BP), molecular function (MF) and cellular component (CC). The results of GO enrichment were verified by the AmiGO database. The Kyoto Encyclopedia of Genes and Genome (KEGG) was used to analyze the biological pathways in which the shared DEPs were mainly involved. The Reactome database was used to verify the results of KEGG. *P*-value <0.05 was deemed as the threshold for the GO enrichment analysis and the pathway analysis. The results were visualized using bubble charts.

### 2.4 Construction of PPI network and identification of hub genes

The protein-protein interactions (PPIs) of shared DEPs were analyzed with an online tool, STRING (v11.5) (34). PPIs with interaction scores >0.4 was considered significant. The results from STRING database were then imported into Cytoscape (v3.9.1) for visualization of the PPIs. A plug-in software, CytoHubba (35), was used to identify the 20 hub genes with eight algorithms (degree, edge percolated component, maximum neighborhood component, maximal clique centrality, closeness, radiality, betweenness, and stress). The intersected hub genes were identified using the upset plot. GeneMania were utilized to construct the co-expression network of intersected hub genes and their related genes (36). Receiver operator characteristics (ROC) curve and the area under the curve (AUC) were utilized to evaluate the diagnostic accuracy of hub genes on COVID-19 and the DTC.

### 2.5 Associations between hub genes and diseases

The associations between the twelve hub genes and related diseases were analyzed using the Comparative Toxicogenomics Database (CTD) (37). CTD is an innovative ecosystem that relates the toxicological information of genes to diseases. Diseases categorized as cancer, respiratory tract disease and viral disease were searched. Bar charts were produced to display the associations between hub genes and COVID-19 and DTC.

## 3 Results

### 3.1 Identification of DEPs in COVID-19 and DTC

A total of 18373 UniProt IDs were converted to 11615 and 6681 gene symbols for the two datasets (IPX0001444000 and IPX0002393000), respectively. By comparing the thyroid samples from patients infected with COVID-19 and those not infected, 137 proteins were confirmed as the COVID-19-DEPs. In addition, 1613 proteins were found to be DTC-DEPs by comparing DTC samples with NTs (Figure 2a&b). The Venn diagram depicted the shared DEPs of two comparisons. Forty-nine common DEPs were identified (Figure 2c & Supplementary data). The expressions of common DEPs in two matrices were displayed with heatmaps (Sup figure 2a&b).

**Figure 2.**
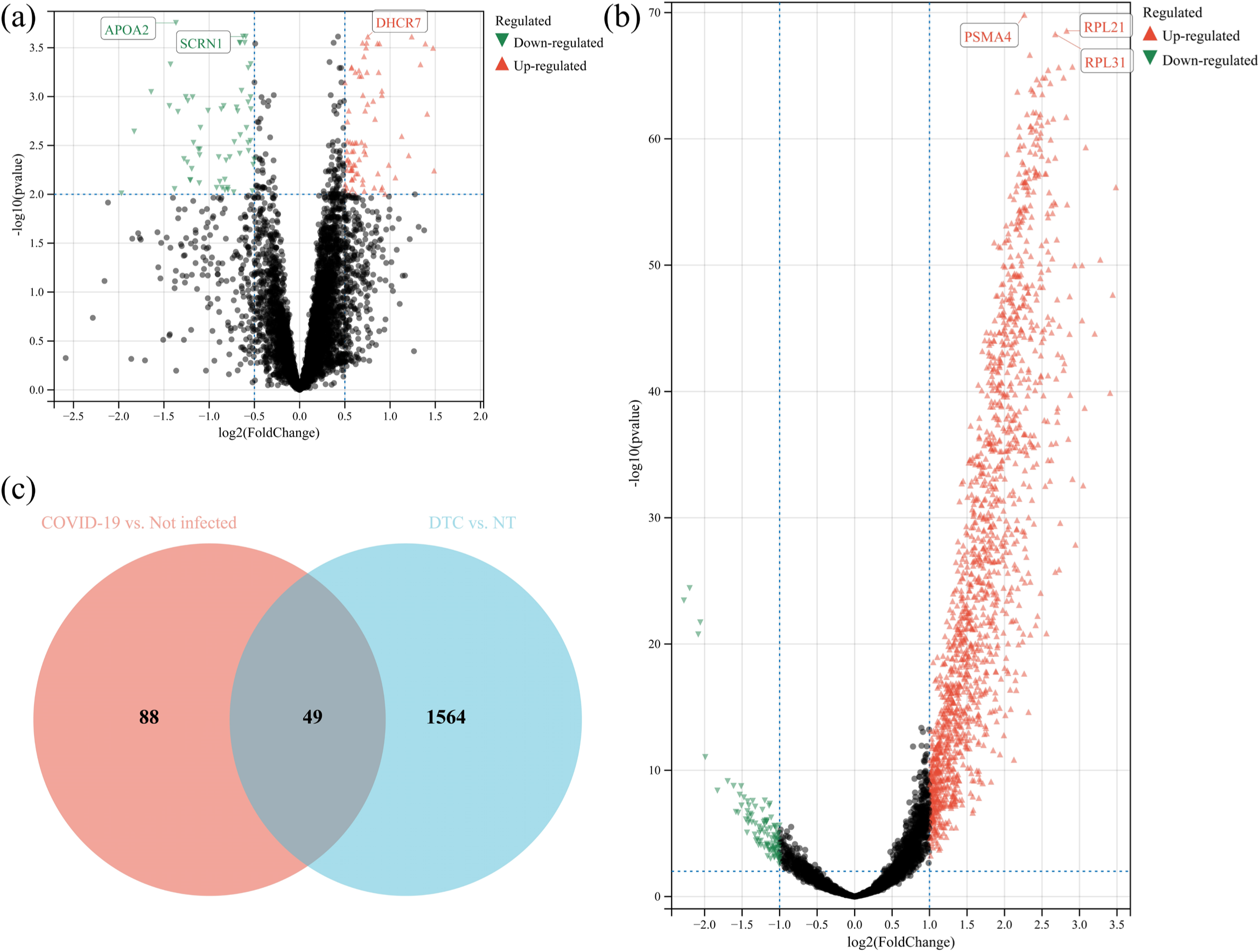
The volcano map of DEPs in two comparisons, (a) COVID-19 vs. not infected, and (b) DTC vs. NT. (c). The Venn diagram of DEPs from two expression matrices. Note: The three DEPs with lowest *p*-value were displayed in the figure. Abbreviations: NT, normal thyroid tissue; DTC, differentiated thyroid cancer.

### 3.2 GO annotation and functional analysis of shared DEPs

GO annotations were performed with the DAVID database. A total of 64 GO terms with *p*-value <0.05 were revealed to be associated with the common DEPs (Figure 3a-c). The most enriched BPs were triglyceride-rich lipoprotein particle remodeling, negative regulation of cholesterol transport and high-density lipoprotein particle clearance. The most enriched CCs were focused on lipoproteins, including spherical high-density, very-low-density and high-density lipoprotein particles. In addition, enzyme inhibitor activity, including lipase, endopeptidase and cysteine-type endopeptidase, ranked top among the enriched MFs. The fold enrichments (FEs) were >30 for the MFs, BPs and CCs mentioned above. The above results were verified with AmiGO database (Supplementary data).

**Figure 3.**
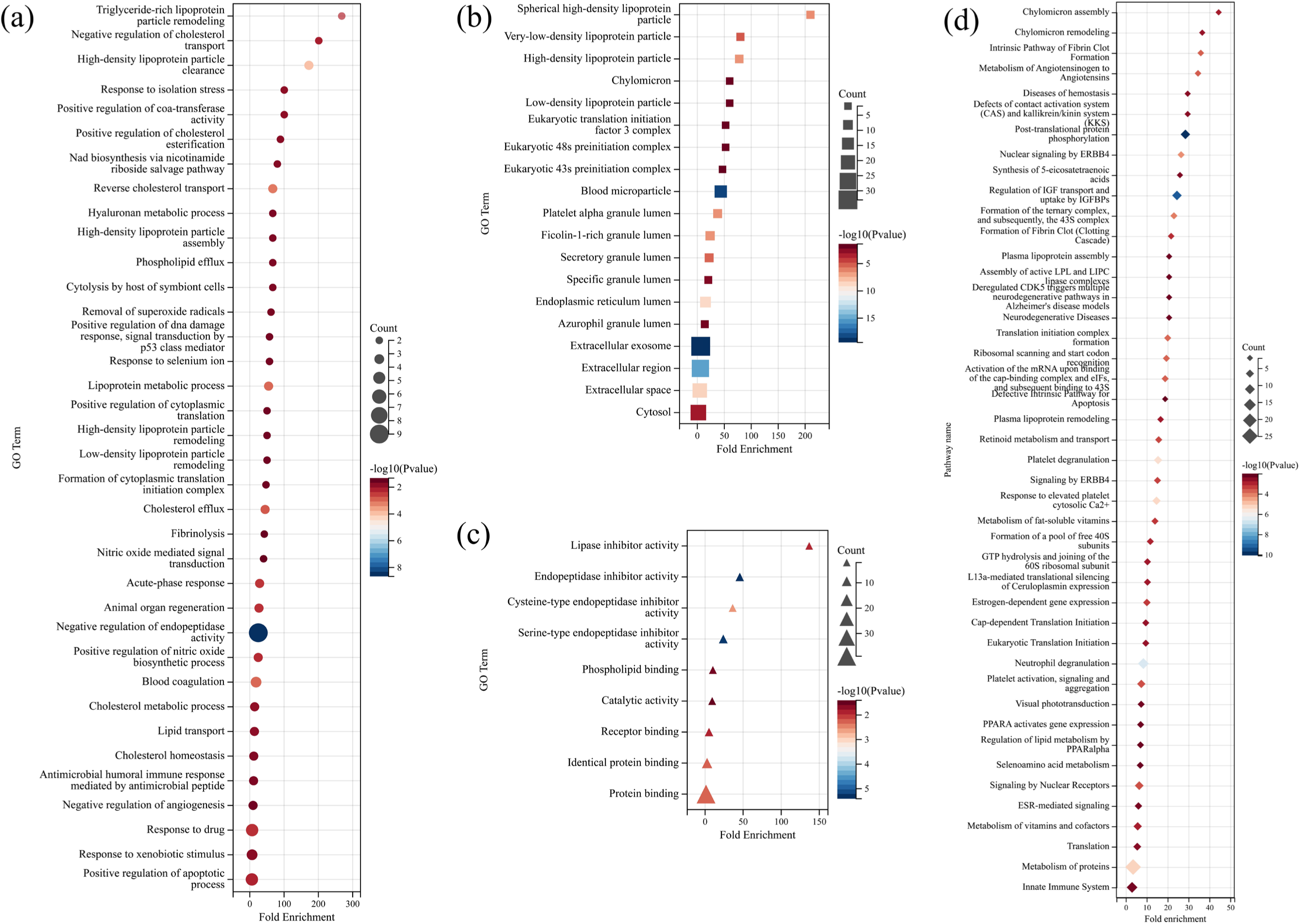
The bubble charts of gene ontology. (a) biological process, (b) cellular component, and (c) molecular function. (d). The results of pathway enrichment analysis. Note: Pathways with *p*-value <0.001 were displayed in the chart, while other pathways could be found in the Supplementary data.

The results of Reactome pathway enrichment analysis were depicted in Figure 3d. The pathways that were most related to common DEPs were chylomicron assembly (FE=44.43), chylomicron remodeling (FE=36.59) and intrinsic pathway of fibrin clot formation (FE=35.89). The pathway analysis from KEGG provided additional information (Supplementary data).

### 3.3 PPI network construction and identification of hub genes

The PPI network included 38 nodes and 149 edges (Figure 4). Eleven common DEPs were excluded due to no connectivity to other genes. Redder color indicated higher connectivity to other genes. Of note, serpin family A member 1 (SERPINA1) was associated with 20 genes in the network.

**Figure 4.**
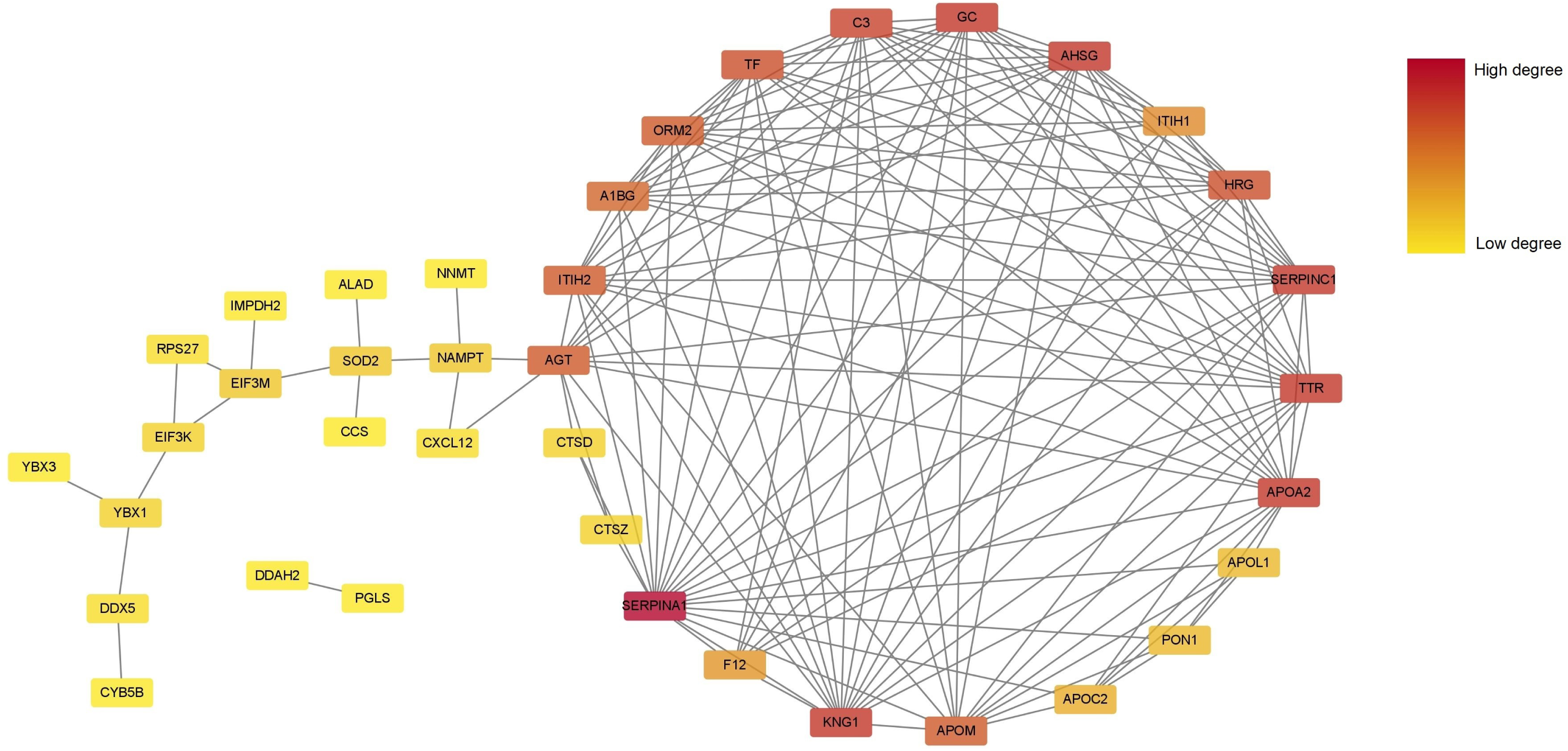
The protein-protein interaction network of shared DEPs. Note: Redder color indicates higher connectivity of the gene with other genes. Disconnected nodes were removed from the diagram.

The top twenty hub gene lists generated by the eight algorithms in CytoHubba were collected (Figure 5a). The intersection of the upset chart identified twelve final hub genes, including AGT, AHSG, APOA2, APOM, C3, GC, ITIH2, KNG1, SERPINA1, SERPINC1, TF and TTR. A complex gene interaction network was constructed with the GeneMania database (Figure 5b). Co-expression contributed to the most interactions in the network (54.09%), followed by co-localizations and physical interactions (21.27% and 19.85%, respectively). The remodeling of protein-lipid complex, plasma lipoprotein particle and protein-containing complex were biological activities most likely associated with hub genes.

**Figure 5.**
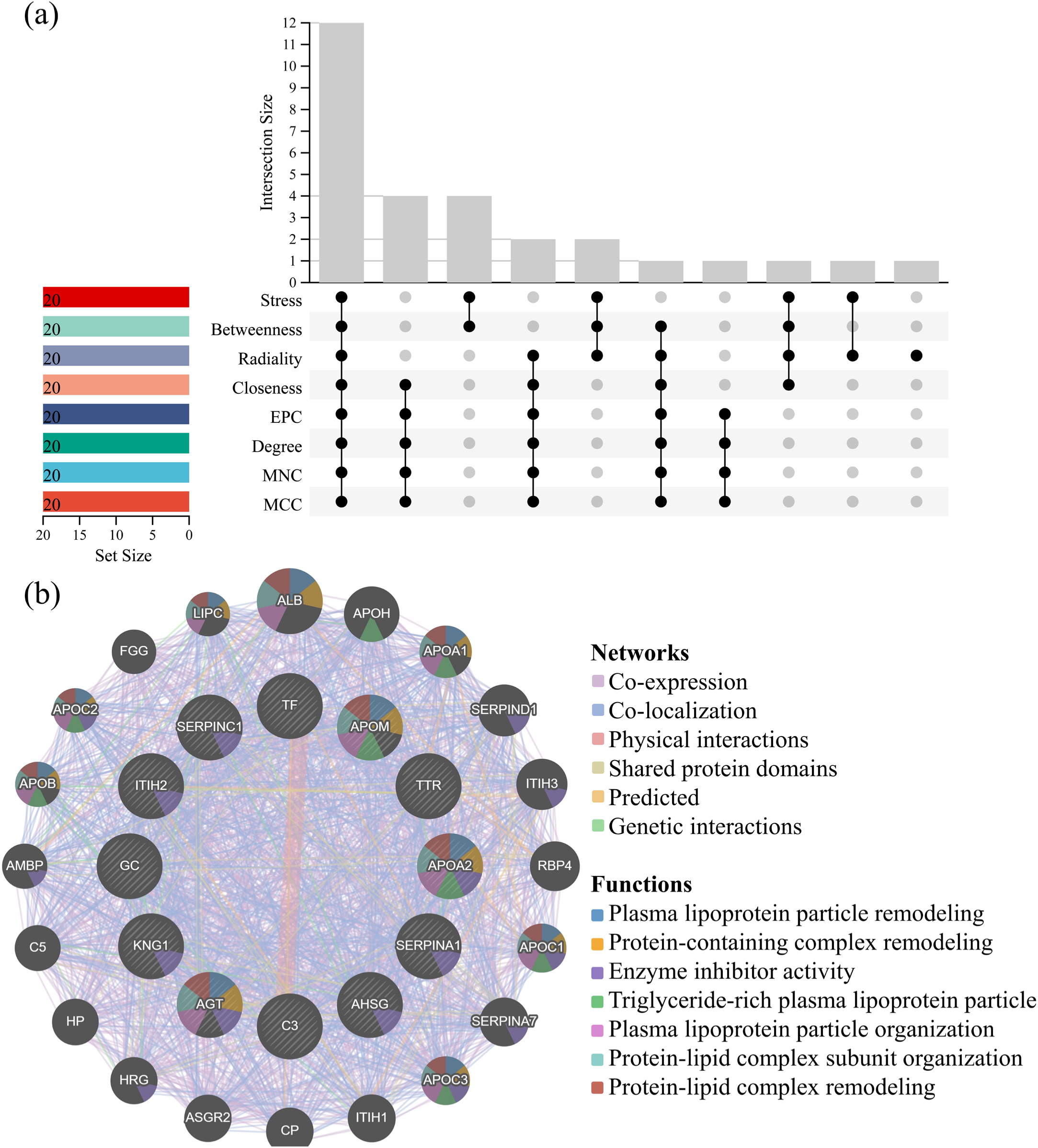
(a) The 12 intersected hub genes identified with the eight algorithms in CytoHubba. (b) The co-expression map of 12 hub genes and their related genes. Twelve hub genes were displayed in the inner circle while the related genes were in the outer circle. Abbreviations: EPC, edge percolated component; MCC, maximal clique centrality; MNC, maximum neighborhood component.

GO annotations and pathway enrichment analysis were conducted based on the twelve hub genes (Sup figure 3a&b). The top five GO terms that related to the hub genes were high-density lipoprotein particle clearance, spherical high-density lipoprotein particle, positive regulation of CoA-transferase activity, positive regulation of cholesterol esterification and high-density lipoprotein particle assembly. After excluding pathways which only one hub gene was involved, Reactome database revealed that intrinsic pathway of fibrin clot formation, post-translational protein phosphorylation and regulation of insulin-like growth factor transport and uptake by insulin-like growth factor binding proteins were the most enriched pathways related to hub genes.

### 3.4 Diagnostic accuracy of hub genes

ROC curves were used to test the diagnostic performance of twelve hub genes (Figure 6a&b). In the COVID-19 dataset, apolipoprotein A2 (APOA2) and serpin family C member 1 (SERPINC1) exhibited the highest diagnostic efficiency (AUC=1.00 for two genes), followed by complement C3 (C3) and inter-alpha-trypsin inhibitor heavy chain 2 (ITIH2) (AUC=0.96 for two genes). In contrast, transferrin (TF) exhibited the best performance (AUC=0.80) in the DTC dataset, followed by transthyretin (TTR, AUC=0.78), alpha 2-Heremans Schmidt glycoprotein (AHSG, AUC=0.78) and vitamin D binding protein (GC, AUC=0.77).

**Figure 6.**
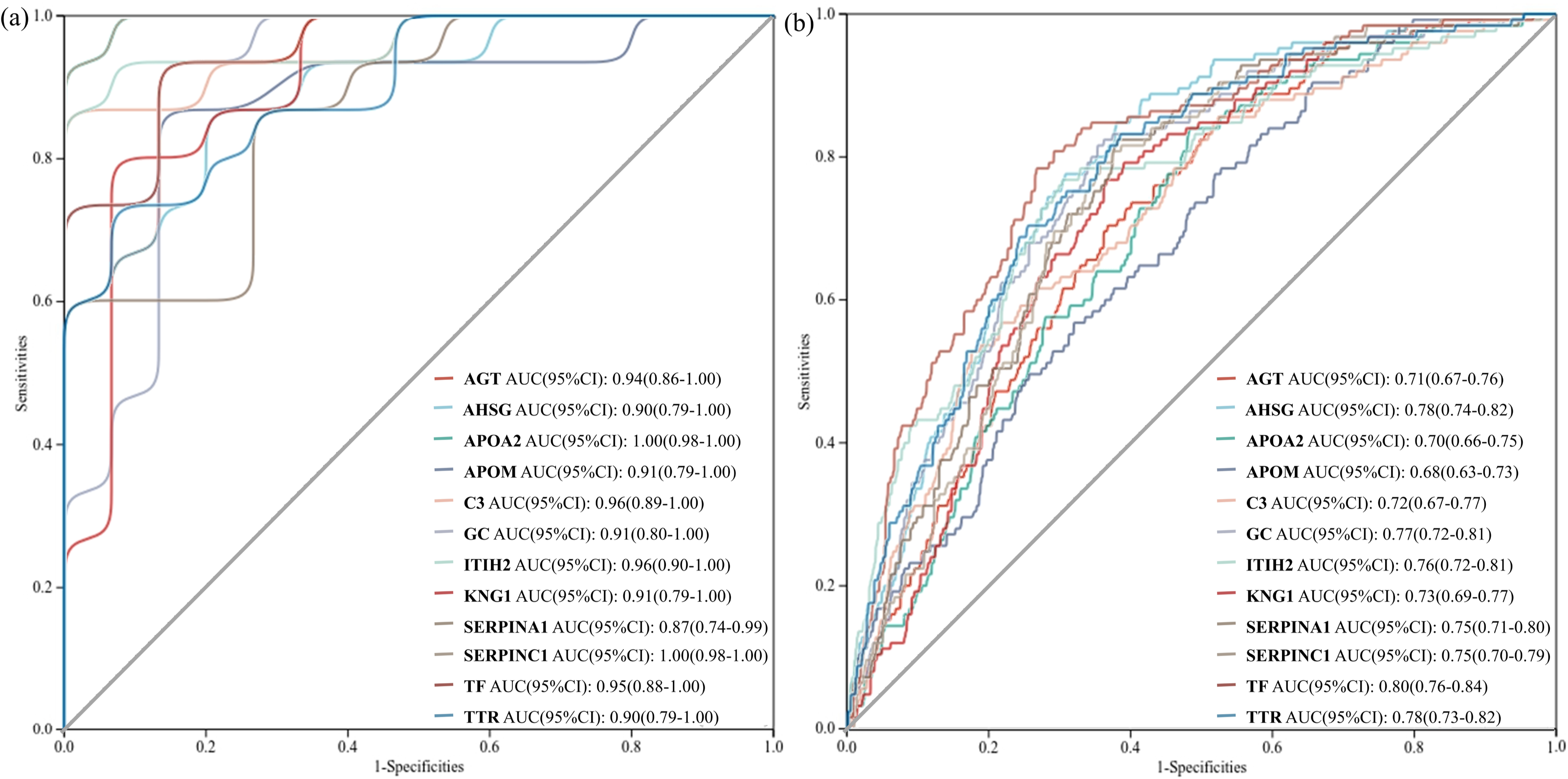
The ROC curve of 12 hub genes in diagnosis of (a) COVID-19 and (b) DTC.

### 3.5 Associations between hub genes and diseases

Ten among the twelve hub genes were shown to have potential interactions with COVID-19 (Figure 7). Notably, direct evidence has proved that angiotensinogen (AGT) participated in the infection of SARS-CoV-2. Besides, C3 and kininogen 1 (KNG1) also had potential interactions with COVID-19. The associations of hub genes and differentiated thyroid cancers were divided into three categories (PTC, FTC and thyroid neoplasms). Transthyretin (TTR), C3, and apolipoprotein M (APOM) exhibited associations with thyroid cancers in all three categories. In addition, ITIH2 had the strongest connection (interference score=8.00) with FTC while TF connected with thyroid neoplasms (interference score=38.25).

**Figure 7.**
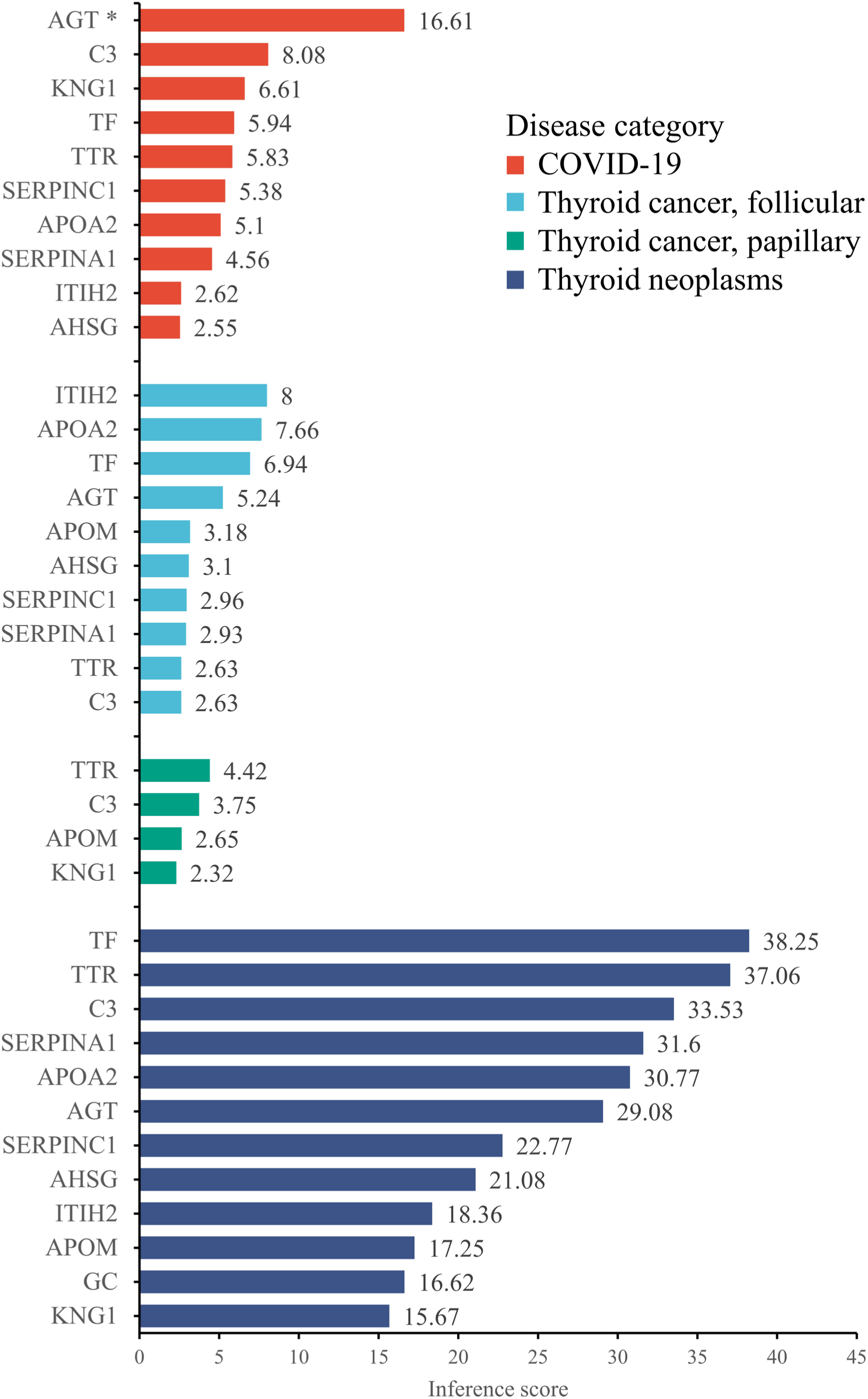
The associations of hub genes and COVID-19 and DTC. * Direct evidence has been revealed between AGT and COVID-19.

## 4 Discussion

In this study, twelve hub genes were identified from 49 common DEPs, including AGT, AHSG, APOA2, APOM, C3, GC, ITIH2, KNG1, SERPINA1, SERPINC1, TF and TTR. GO annotations and pathway enrichment analysis revealed several biological activities that might involve in the interaction of COVID-19 and DTC. Briefly, the *in vivo* transport and metabolism of lipid and cholesterol are supposed to be the most likely common pathogenic mechanism in COVID-19 and DTC. In addition, vasodilation and coagulation process may also play roles, whereas chronic inflammation and immune response showed little effects.

The pathogenic effects of COVID-19 on thyroid gland have been gradually noticed since the outbreak of the pandemic in Wuhan. In March 2020, a retrospective clinical study in Wuhan evaluated the clinical and laboratory data of 274 patient infected with the new coronavirus disease (38). In comparison with those who finally recovered from COVID-19, deceased cases presented with lower levels of serum thyroxine and triiodothyronine. Confusingly, the thyroid stimulation hormones were also decreased in these patients. Subsequent studies verified that ACE2 is the main host cell receptor for SARS-CoV-2 (39). Taken that ACE2 is widely expressed in multiple organs, including thyroid and pituitary (25), we may hypothesized that the hypothalamus-pituitary-thyroid axis is impaired during the infection. In comparison with healthy controls, infected patients were more likely to develop hyperthyroidism, as revealed by a retrospective study (40). Another study recruited COVID-19 patients in the intensive care units (41). The incidence of subacute and atypical thyroiditis was much higher in those with high intensity of care (10% *vs.* 0.5%). Besides, Grave’s disease could also be triggered by SARS-CoV-2 infection in patients with or without a previous Grave’s disease history (42, 43). Direct viral infection and hyperinflammatory state by COVID-19 are both believed to induce thyroid autoimmune disorders.

Although numerous evidences have proven the damage of coronavirus on thyroid tissue, the linkage between thyroid cancer and COVID-19 is unclear. Our previous study in Wuhan showed that thyroid cancer behaviors were generally more aggressive compared to those before the pandemic (22). More cases with multiple lesions, extrathyroidal extension and lymph node metastasis were discovered. A multicenter cohort study in Los Angeles suggested that comorbidities and old age are hazard factors for patients diagnosed with both thyroid cancer and COVID-19 (21). Another nationwide study evaluated the COVID-19 outcomes and mortality in thyroid cancer patients with COVID-19 infection (44). No elevation in mortality and morbidity was revealed. The conclusions were verified by a multicenter cohort study conducted in South Korea (45). Nevertheless, considering the fact that a small proportion of patients receive tyrosine kinase inhibitor therapy or have distant metastasis, it is reasonable to anticipate that these patients are at high risk for complications of COVID-19.

Among the twelve hub genes identified, AGT is the gene most related to COVID-19. AGT gene encodes for the angiotensinogen precursor, which is quickly converted to angiotensin I with the regulation of blood pressure and renin. Two retrospective studies drew the same conclusion that AGT rs699 C>T single-nucleotide polymorphism could increase the risk and severity of COVID-19 (46, 47). Another study collected serum samples from 45 infected patients and 34 controls (48). Proteomic analysis revealed that AGT was elevated in almost all infected patients and its serum concentration was strongly indicative of disease severity. Taken that ACE2 is the key factor in the internalization of SARS-CoV-2, the renin-angiotensin-aldosterone (RAS) system is supposed to over-activate in response to COVID-19. The RAS system was less studied in thyroid carcinomas. A clinical study revealed a significantly increased expression of ACE2 in cancer tissues in comparison with benign nodules (49). The result suggested that the RAS system might involve in tumor progression, as supported in previous basic research (50). However, direct and solid evidences are lacking in the relationship between AGT and thyroid cancers.

KNG1 is an essential factor in the kallikrein-kinin system and plays an important role in blood coagulation. The over expression of KNG1 could inhibit proliferation and induce apoptosis of glioma cells (51). Its anticancer mechanisms include the induction of G1 phase cell cycle, the activation of caspase 3 and 9 and the inhibition of vascular endothelial growth factor expression. Of interest, the expression of KNG1 was increased in colorectal cancers (52, 53). The phenomenon could be explained by the compensative effect of cancer tissues. In patients with COVID-19, KNG1 was shown to be upregulated in circulation together with other regulators of coagulation (plasminogen, fibronectin 1, etc.) (54), suggesting that coagulation dysfunction might occur.

Lipid and cholesterol metabolism and transport are biological processes most likely involved in the common pathogenic mechanism of COVID-19 and DTC. Schmelter *et al.* collected 482 serum samples from infected patients and healthy volunteers and tested the serum lipid profile. They discovered that very-low-density lipoprotein, intermediate-density lipoprotein particles and associated apolipoprotein B were significantly elevated in patients with SARS-CoV-2 infection, whereas cholesterol and APOA2 were decreased (55). In another study using the proteomic approach with 41 patients, APOM in high-density lipoprotein was inversely associated with the odds of death due to COVID-19 complications (56). Since numerous studies have reported dyslipidemia due to COVID-19, recent researches uncovered the underlying mechanisms. When SARS-CoV-2 enters the host cell via the receptor ACE2, cholesterol is essential for the formation of syncytia (57). This might help to explain the success of lipid apheresis as the treatment for patients with long-COVID syndromes (58). Previous studies have proved that dyslipidemia, especially hyperlipidemia and hypercholesterolemia, could increase the risk of various cancers (59, 60). Excess lipid and cholesterol would increase intracellular oxidative stress and consequently promote carcinogenesis and aggressiveness (61). Evidence has shown that lipid metabolism was impaired in those individuals with long-COVID-19 (62), we could postulate that dyslipidemia due to post-acute sequelae of COVID-19 might elevate the risk of thyroid cancer in a long term.

The associations between other hub genes and COVID-19 and DTC were also studied. C3 encodes for complement component 3. The complement system can be activated by several different mechanisms in acute inflammation and malignancies. The clinical studies of C3 in COVID-19 are controversial. Some reports favored that decreased C3 levels were associated with the disease severity (63, 64), while another research found no statistical significance (65). A clinical study evaluated the proteomic profile in fine needle aspiration biopsy (66). C3 expression was discovered to decrease in PTC tissues, which could activate the downstream complement system and exert a compensative anticancer effect. Similar to this article, GC expression was also altered in our study. A previous research observed a decreased GC in PTC tissue was related to enhanced tumor proliferation and migration (67). The link between COVID-19 and GC is unclear currently. A commentary postulated that GC might play a role in vitamin D deficiency in infected patients (68). Besides, TF, an iron-related serum protein associated with chronic inflammation, was proved to be indicative of the severity and coagulative state in COVID-19 patients (69–71). However, the association between TF and DTC, if exists, was unknown. Of the rest five hub genes, the levels of AHSG, ITIH2 and SERPINA1 were found to be the predictors of severity in patients with SARS-CoV-2 infection (72–74). Besides, the expressions of AHSG, SERPINA1 and TTR were altered in PTCs (75–77). Of note, the above researches utilized omics methods to evaluate the expression profiles but the underlying mechanisms between molecules and diseases were not explored. The roles of SERPINC1 in COVID-19 and thyroid cancer have not been studied.

One letter published in Sept 2022 conducted a bioinformatic analysis similar to our study (78). In this study, a database from iProx was analyzed for the identification of DEPs. The transcription profiles of thyroid cancers and normal controls were downloaded from the data from the Cancer Genome Atlas (TCGA) database. Using a different algorithm and criteria (*p*<0.05, log2|FC|>1), this research identified ten more DEPs for COVID-19-infected thyroid tissues (137 in our study and 147 in this study). Besides, 31 differentially expressed genes were determined with 568 TCGA samples. However, only one common gene/protein (coagulation factor XII) was identified with the Venn diagram. This pilot research has some defects in the study design. First, samples in TCGA databases were collected from different sources and batches. The batch effects between arrays were not normalized before analysis. Second, differential expression analysis was conducted on both mRNA and protein levels. Altered mRNA expressions do not certainly indicate the changes of translation, and vice versa. Finally, the GO annotations and pathway enrichment analysis were not conducted, which limited the generalization of conclusions.

There are some limitations in our study. First, due to the lack of patients with concomitant thyroid cancer and COVID-19, the results of our study were not validated in human samples. Second, we identified some novel molecules associated with SARS-CoV-2 infection or DTC, the underlying mechanisms were not evaluated with *in vitro* or *in vivo* experiments. Besides, only 30 samples were extracted and analyzed in the matrix of COVID-19. The sample size was relatively small.

In conclusion, our study identified twelve hub genes from 49 common DEPs. GO annotations and pathway enrichment analysis focused on lipid and cholesterol metabolism and transport, followed by vasodilation and coagulation process. The common pathogenic mechanisms suggest the infection of SARS-CoV-2 might exert adverse effects on patients with DTCs. Further clinical and basic researches are needed to verify our conclusions.

## Data Availability

The two proteomic profiles can be downloaded from iProx database (IPX0001444000 & IPX0002393000). Supplementary figures and data files were deposited in Mendeley database (DOI: 10.17632/5cg2dmz5nb.2).

https://www.iprox.cn//page/SCV017.html?query=IPX0001444000

https://www.iprox.cn//page/SCV017.html?query=IPX0002393000

https://data.mendeley.com/datasets/5cg2dmz5nb/2

## Abbreviations

ACE2: angiotensin-converting enzyme 2
AGT: angiotensinogen
AHSG: alpha 2-Heremans Schmidt glycoprotein
APOA2: apolipoprotein A2
APOM: apolipoprotein M
AUC: the area under the curve
BP: biological process
C3: complement C3
CC: cellular component
COIVD-19: coronavirus disease 2019
CTD: Comparative Toxicogenomics Database
DEP: differentially expressed protein
DTC: differentiated thyroid cancer
EBV: Epstein-Barr virus
FC: fold change
FE: fold enrichment
FTC: follicular thyroid cancer
GC: vitamin D binding protein
GO: Gene Ontology
ITIH2: inter-alpha-trypsin inhibitor heavy chain 2
KEGG: Kyoto Encyclopedia of Genes and Genome
KNG1: kininogen 1
MF: molecular function
NT: normal thyroid tissues
PPI: protein-protein interaction
PTC: papillary thyroid cancer
RAS: renin-angiotensin-aldosterone
ROC: receiver operator characteristics
SARS-CoV-2: severe acute respiratory syndrome coronavirus 2
SERPINA1: serpin family A member 1
SERPINC1: serpin family C member 1
TCGA: the Cancer Genome Atlas
TF: transferrin
TriC/CCT: T-complex protein 1 ring complex/chaperonin-containing T-complex protein 1
TTR: transthyretin.

## Disclosures

### Authors contribution

Conceptualization: HQ. Liu & C. Chen

Methodology: HQ. Liu, JX. Wang & C. Chen

Formal analysis: HQ. Liu, JX. Wang & D. Yang

Investigation: HQ. Liu, JX. Wang & D. Yang

Writing – original draft preparation: HQ. Liu & JX.

Wang Writing – review and editing: D. Yang & C. Chen

Supervision: C. Chen

Funding acquisition: C. Chen

## Acknowledgments

We wish to thank researchers who provide the two proteomic profiles.This article is dedicated to the memory of those who lost their lives during the pandemic.

## Data availability

The two proteomic profiles can be downloaded from iProx database. No new data was generated in this study.

