## Supplementary figures for "Common pathogenic mechanisms for COVID-19 and differentiated thyroid cancer: a proteomic analysis"

Supplementary Figure 1. The boxplots and density diagrams of protein expression distribution in two matrices.

Supplementary Figure 2. The heatmaps of common DEPs in the two expression profiles. (a) COVID-19 compared with not infected, (b) DTC compared with NT.
Note: Euclidean distance and Complete method were used in the clustering. Z-scores were calculated.

Supplementary Figure 3. The bubble chart of (a) gene ontology and (b) pathway enrichment analysis for hub genes.

Supplementary figure 1

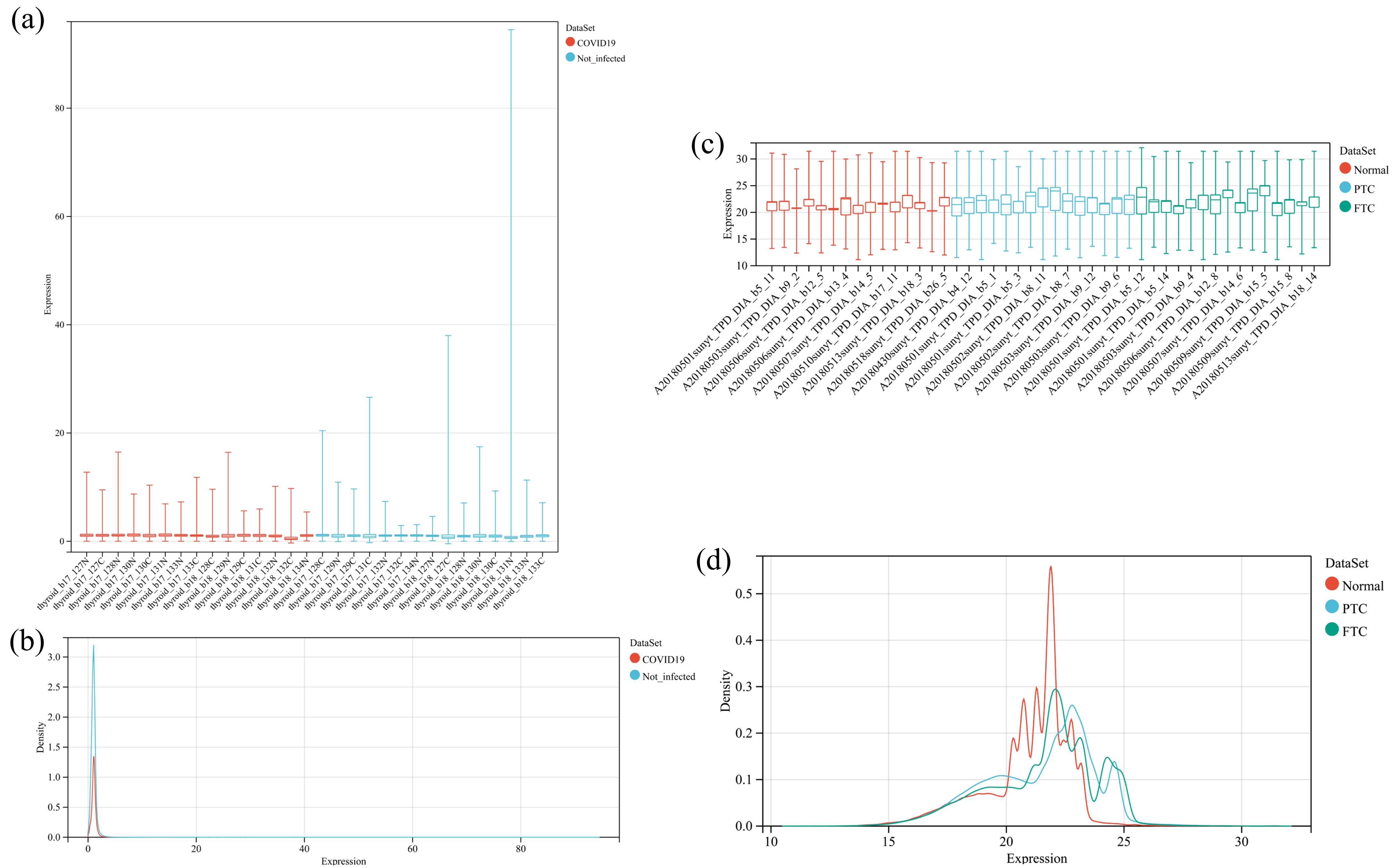

Supplementary figure 2

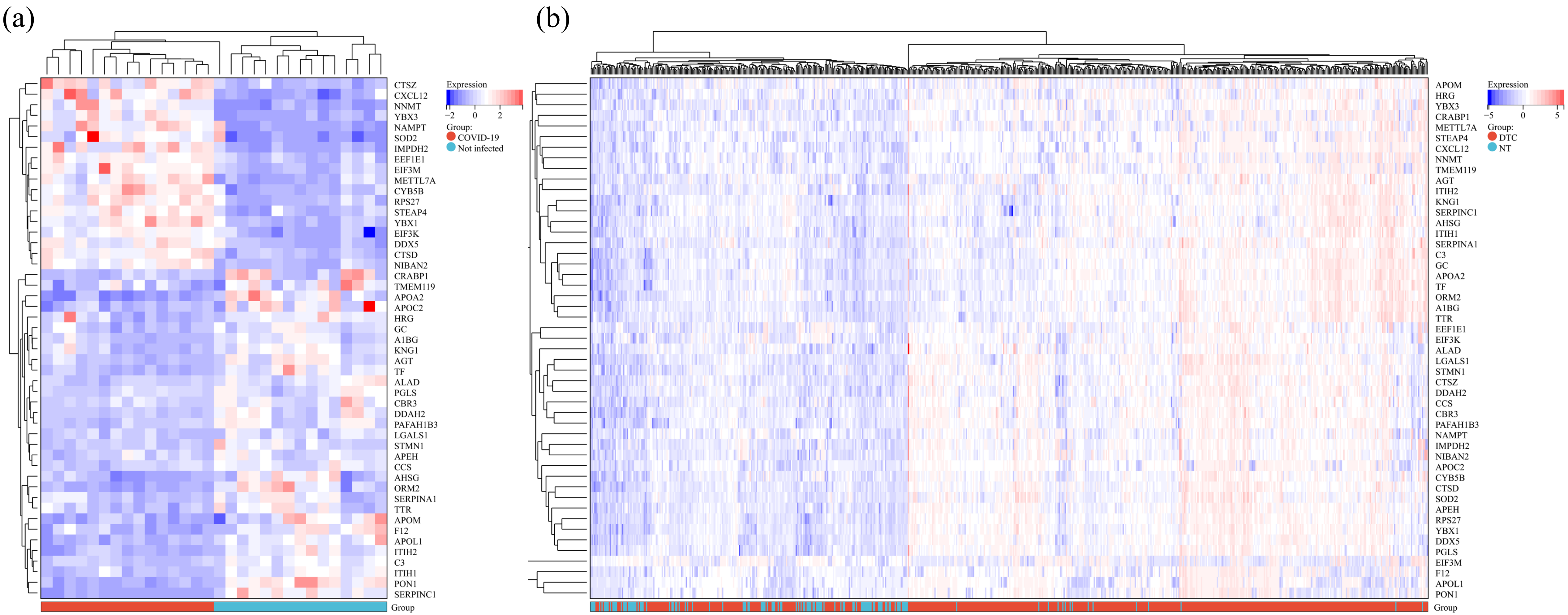

Supplementary figure 3

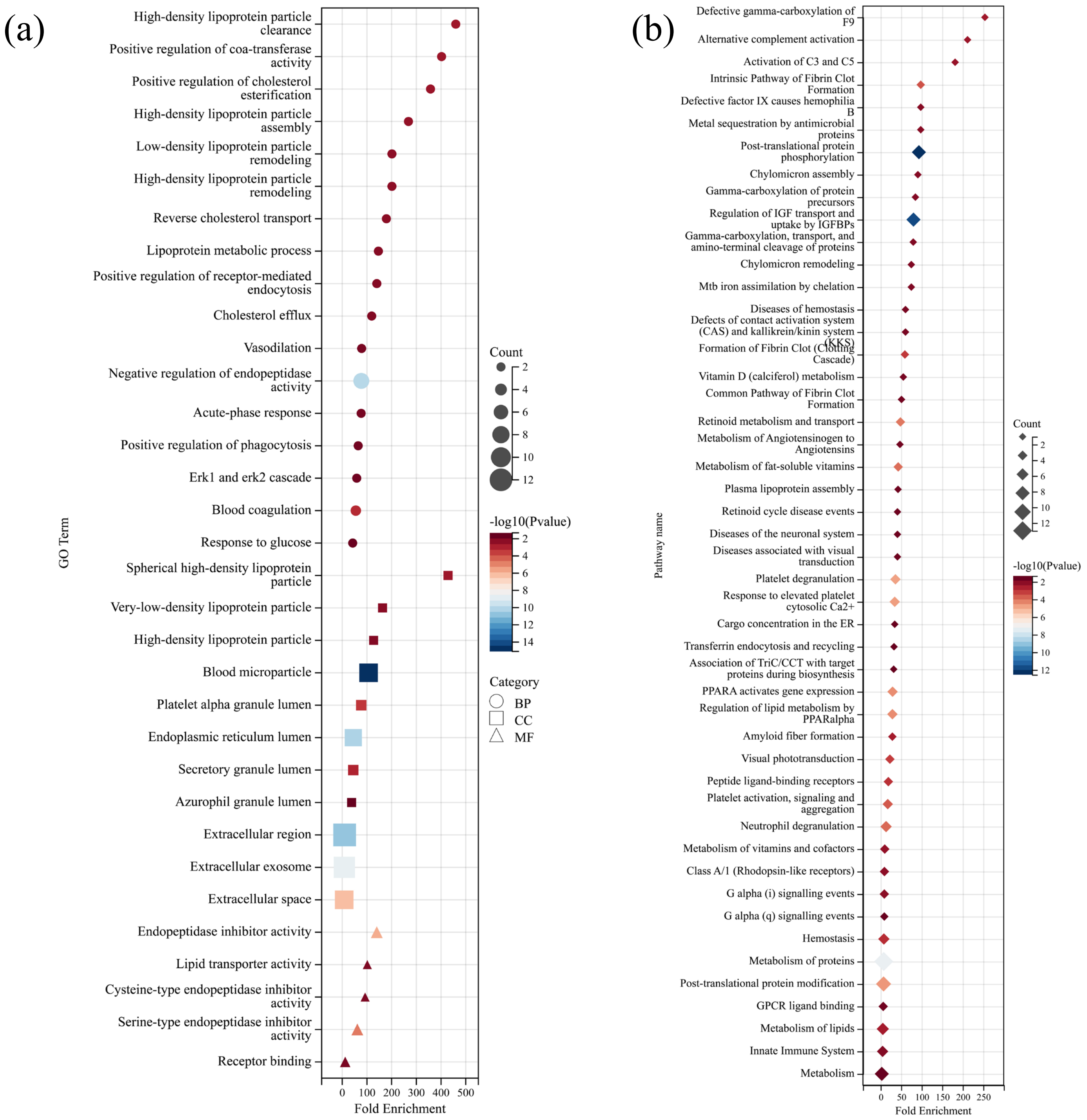
